# Predictors of perinatal mortality in the seven major hospitals of Lusaka Zambia: A Case Control Study

**DOI:** 10.1101/2024.05.21.24307685

**Authors:** Musonda Makasa, Patrick Kaonga, Choolwe Jacobs, Mpundu Makasa, Bellington Vwalika

**Affiliations:** University of Zambia School of Public Health, Department of Epidemiology and Biostatistics, Lusaka, Zambia; University Teaching Hospital, Women and Newborn Hospital, Lusaka, Zambia; University of Zambia School of Public Health Department of Community and Family Medicine Lusaka, Zambia; University of Zambia School of Medicine Department of Obstetrics and Gynaecology Lusaka, Zambia

**Keywords:** Perinatal, proportions, predictors, mortality

## Abstract

**Background:** Over 2.6 million babies are lost later in pregnancy, during labour, and or in the first week of life. Global perinatal mortality reduced from 5.7 million since 2000 to 4.1 million in 2015. High-income countries account for 45% of this data. The rest are in low-income countries, 77% of which are in sub-Saharan Africa. Perinatal mortality rates for sub-Saharan Africa and Zambia are 42.95 and 33/1000 live births, respectively. The aim of this study was to determine the predictors of perinatal mortality at the seven major hospitals of Lusaka, Zambia.

**Methods:** This was a multi- centre unmatched case control study from September 2023 to January 2024. Cases included perinatal death (>24 weeks gestation or >500g stillborn, and death of neonate within seven days of life) and controls were live births. Stepwise multivariate logistic regression analysis determined predictors using adjusted odd ratios and p-values.

**Results:** The study had 630 participants, 210 cases and 420 controls were analysed: ratio 1:2. Antenatal care booking after 12 weeks gestation had almost three times odds of experiencing perinatal (AOR 2.909, 95% CI: 1.97-4.296), p <0.001 compared to those who booked early. Walking as means of reaching healthcare facility had over three odds perinatal mortality (AOR3.482, 95% CI: 1.87-6.49) than personal transport users. Anaemia during pregnancy had over three times risk of perinatal death (AOR 3.581, 95% CI: 1.72- 7.44) than those without it. History of loss of baby before birth had five fold odds to experience perinatal mortality than to those who had not (AOR 5.047, 95% CI: 2.99-8.51).

**Conclusion:** This study revealed that late antenatal care booking, walking, as means of transport to access health facility, anaemia in pregnancy, and previous history of loss of baby before birth perinatal death were the main predictors with statistical significance of perinatal death experience.

## Introduction

Perinatal mortality remains a significant global health challenge particularly in low-income settings (1). An estimated 2.6 million babies are lost per year during pregnancy, labour, and or in the first seven days of new-borns’ lives. Attempts to avoid these perinatal deaths have not yielded much. For example, until recently most stillbirths not accounted for in the worldwide data tracking, while social recognition and lack of investment and programmatic action have contributed to this problem (2). The Maternal Mortality Estimated Interagency Group (MMEIG) reported an estimated 295,000 maternal deaths globally in 2017, and 196,000 (66%) were from sub-Saharan Africa (SSA) (3). In low-income settings, in over a third of these maternal deaths, half of the stillbirths and almost a quarter of neonatal deaths occur before and during childbirth (4, 5). Literature shows a strong linkage between maternal deaths and perinatal mortality because for every maternal death, there is an estimated 10 perinatal deaths. Almost two thirds of maternal death causes also account for the causes of perinatal deaths (6). By definition, perinatal mortality is the loss of a foetus after 24 weeks of gestation; birthweight of ≥500g; and loss of newly born within 7 days of life. Whereas early neonatal mortality is a subset of perinatal mortality and refers to loss of newly born within 7 days of life (7) and for purposes epidemiological studies and statistics (8).

The World Health Organization Sustainable Development Goal (SDG) number 3.2 targets to end preventable neonatal deaths by reducing neonatal mortality rate to at least 12 per 1000 live births by 2030 (1). The global perinatal mortality reduced from 5.7 million since the launch of the Millennium Development Goals in 2000 to 4.1 million by the time of the launch of the SDG (9). Similarly, the global stillbirth rate also declined from 24.7 in 2000 to 18.4 per 1000 live births in 2015 (10). The neonatal mortality rate also demonstrated a downward trend from 37 in 1990 to 19 deaths per 1000 live births around 2016 (11). These strides however, have only been realistic in High-Income Countries (HIC) that account for only approximately 45% of the global records on perinatal mortality. SSA has the highest Perinatal Mortality Rate (PMR) (42.95 per 1000 live births) with Nigeria leading followed by Ethiopian 40.9 and 49 per 1000 live births respectively, whereas southern Africa is approximately 30.3 according to systematic reviews and meta-analyses by Akombi and Renzaho (12), (13).

Zambia is not an exception to the above-mentioned protracted decline of perinatal mortality figures. The PMR for Zambia is 33 per 1000 population (14), which is above the SSA average and therefore still far from attaining the SDG and Vision 2030 set targets (15). This study’s aim was to assess the predictors of perinatal death in Lusaka urban district of Zambia, which is the most populated city in Zambia and has the highest birth rate. Additionally, the study also aimed to provide evidence to that guide program interventions to reduce the PMR in this setting.

## Materials and Methods

This was a multicentre study at the seven major hospitals of Lusaka urban district of Zambia. The data collection process started in September on the 9^th^ day of 2023 and concluded in January the 31^st^ 2024. The study did not involve minors. Whereas, the participants were prospectively recruited as and when cases occurred and if agreeable to partake of the research process. Information about the research was share with the participants through the participant information sheet before obtaining an informed written consent. Data collection was through an interviewer-structured questionnaire. They included two tertiary and five first level hospitals namely the Women and Newborn and Levy Mwanawasa University Teaching Hospitals, Chawama, Chilenje, Chipata, Kanyama, and Matero first level hospitals. In accordance with the 2022 census report, the Lusaka is the most populated city in Zambia and has female population of 1,590,922 with a national fertility rate of 4.4 (16). These facilities also serve the most densely populated populations in the country. The table 1.0 hereafter shows the distribution and total number of deliveries from all study facilities during period January to December 2023.

**Table 1.0.**
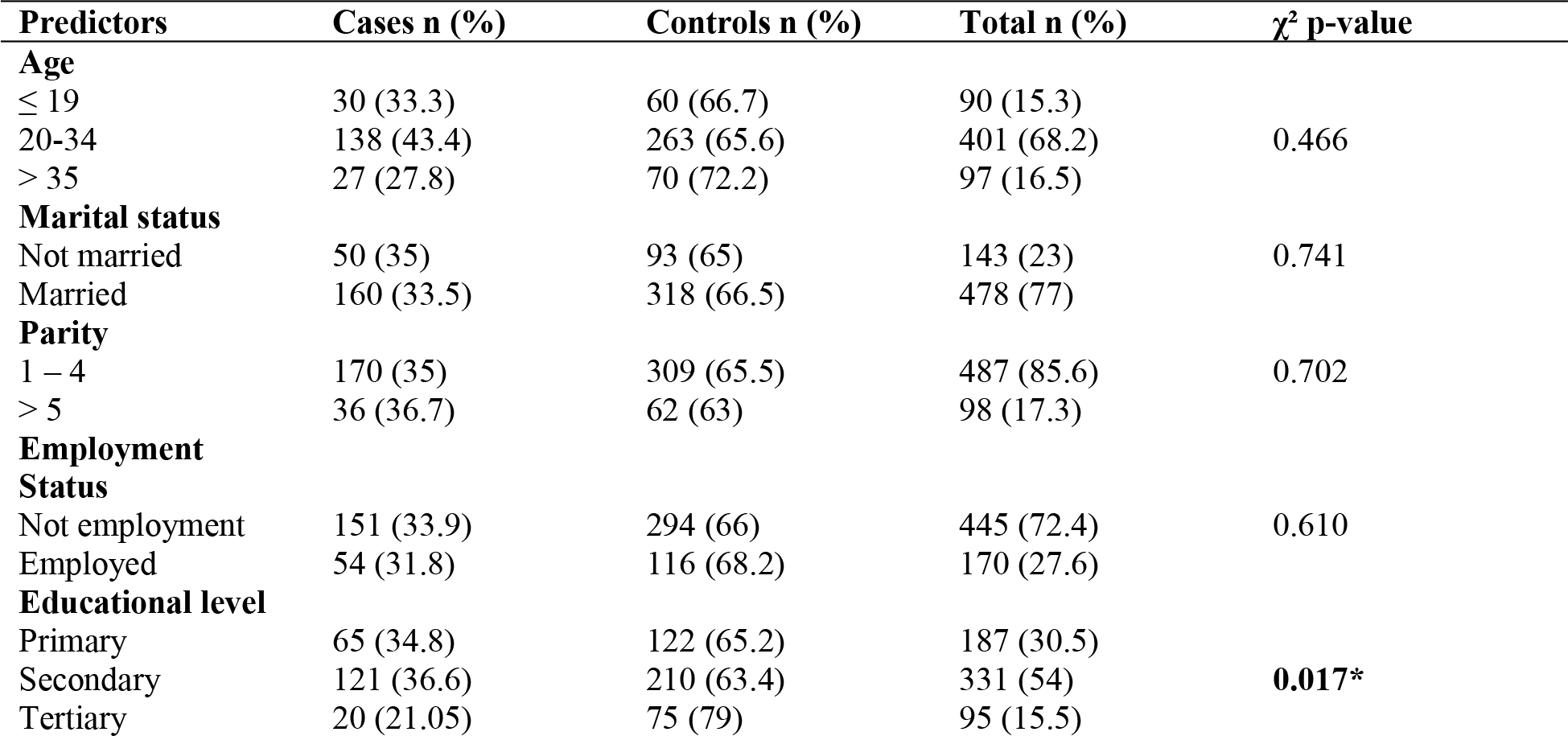
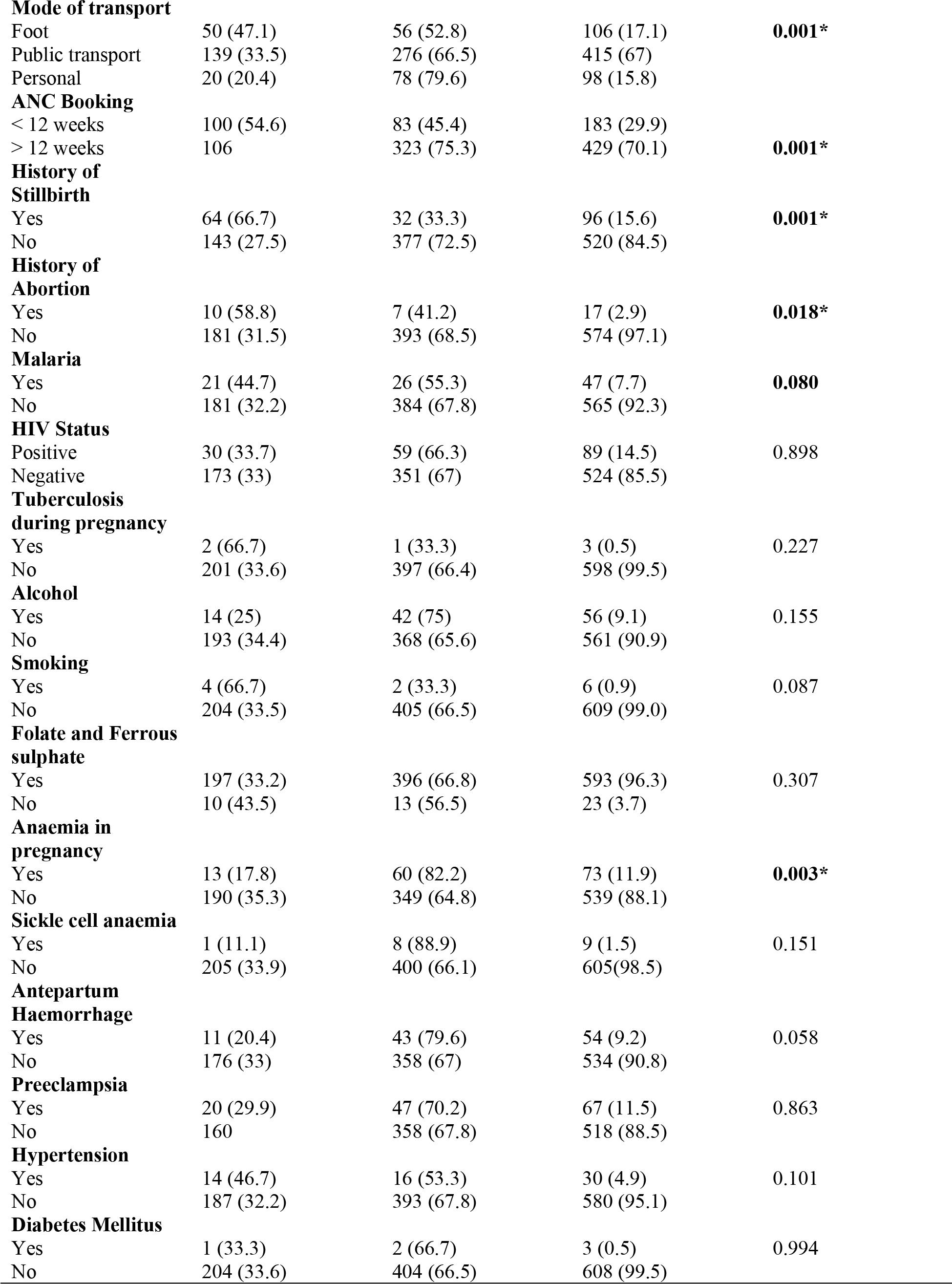
Summary of the maternal socio-demographic characteristics frequency distribution.

### Study design

This was an unmatched case-control study with a ratio of 1:2 aimed at measuring the proportions and determination of predictors of perinatal mortality at the selected study site. Purposeful sampling for the cases if they met the inclusion criteria was used due to the rarity of the outcome. Cases were defined as stillbirth above 24 weeks of gestation and early neonatal deaths within seven days of life from from the study sites. Whereas, two controls for every case underwent systematic random sampling by selecting women that had a live birth before or after incidence of a case. For early neonatal deaths controls were those who delivered at least within 24hrs of the date of birth of of the this case. After sharing the participant information sheet and obtaining an informed consent. Files were reviewed followed by interviewer administered collection of data using a standardized structured questionnaire on a google sheet that was managed by the Principal Investigator (PI). The google data collection sheet was monitored on a daily basis by the PI.

### Study population

Study population involved women from 18 years old and above who sought and had childbirth services at the study sites and met the inclusion criteria. Refer to the following figure 1.0 flowchart that illustrates the source of the sample, eligibility screening process and how the final sample was arrived at.

**Figure 1.0.**
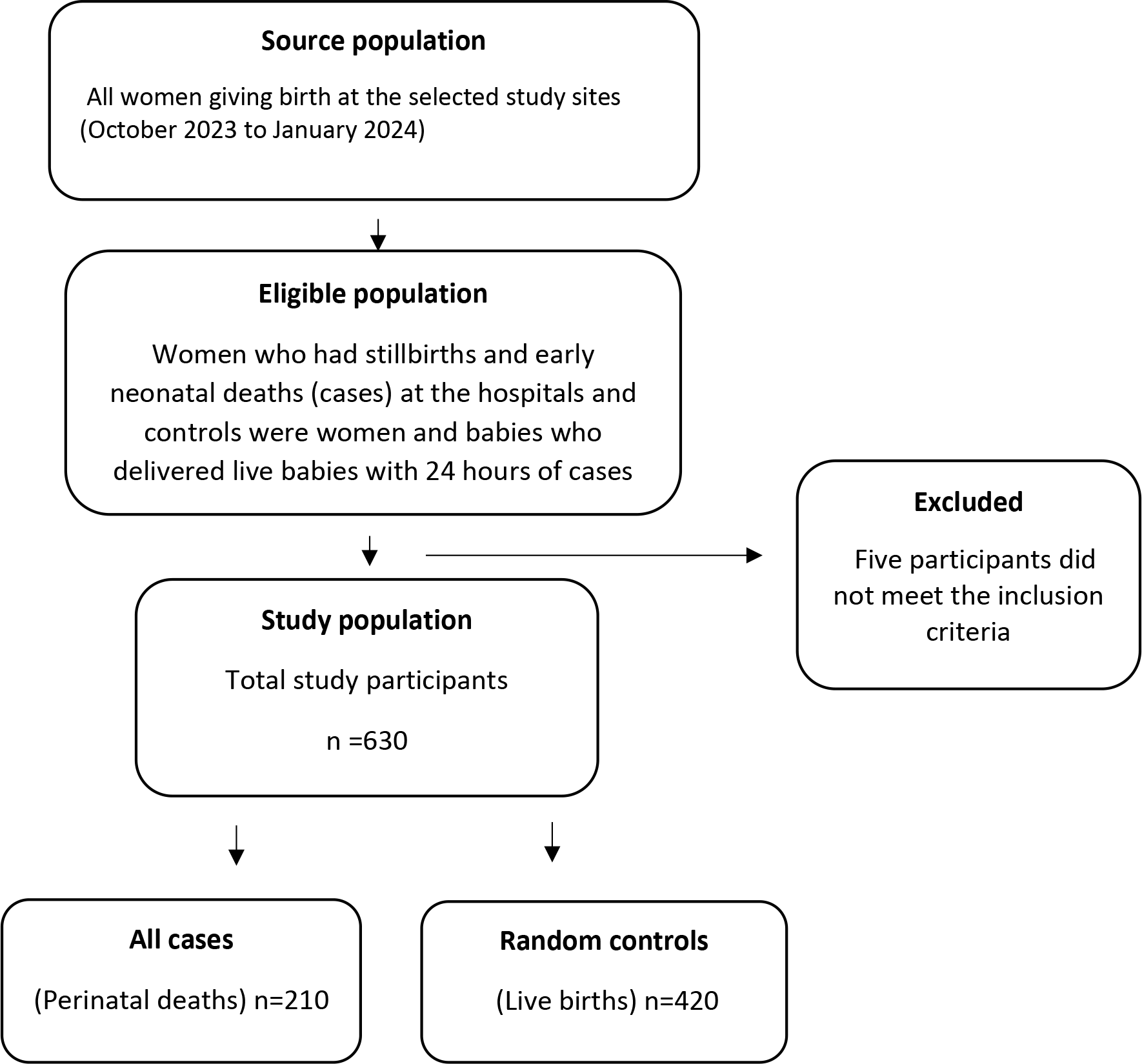
Selection of study participants’ flow chart

### Inclusion criteria

Cases included stillbirths with mothers above 18 years of age that were delivered with a gestational age above 24 weeks of gestation. In the absence of gestational age a birthweight of ≥500g was eligible for purposes of this study. Early neonatal deaths eligibility was mortality that occurred within seven days of birth. Controls included women also above 18 years of age with live babies recruited within 24 hours of a case.

### Exclusion criteria

Women who chose to withdraw from the study, below the age of 18 as they could not sign for consent. Any pregnancy below 24 weeks of gestation or in the absence of gestational age birthweight less than 500g. Women who chose not to participate after sharing information from the participant information sheet.

### Sample size calculated

The sample size for the study was calculated based on the assumptions that two controls per care (r = 2), with 80% power of the study (type II error 20%, i.e. β = 0.2), confidence interval of 95% with 5% type I error (i.e. α = 0.05). With an assumed odds ratio of 2.5 for differences based on findings in previous similar studies. They reported for example associated potential risk factors included extremes of birthweight, post term delivery, infections, haemorrhage and previous history or perinatal mortality (17, 18). And using Zambia’s PMR of 33 per 1000 live births (14), the following formula by Charan and Biswas (19) was used:

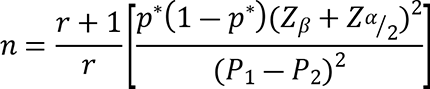

Where *n* = is the sample size, *p*^∗^average proportion exposed which was given by the sum of proportion exposed cases (*P*_1_) and proportion of control *P*_2_ divided by 2 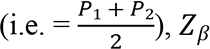, is the standard normal value for power (for power of 80%, *Z*_*β*_ = 0.84), *Z*_*α*/2_ is the standard normal value for the level of significance which is usually given as α = 0.05 and it’s value is 1.96, and r is the ratio of controls to cases, in our case the ratio is 2 to 1 which means our r = 2; To find P_2_, we use the formula:

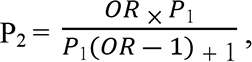 here OR is the odds ratio and P_1_ is the proportion with a specific risk factor in the control participants. Assuming OR = 3.2 based on pooled analysis with strong association of perinatal mortality due to lack of antenatal care (6): P_1_=0.033 in the formula above we get the value of P_2_ as 0.09845

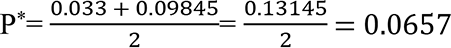

Replacing the above information in the prescribed formula above the following was the targeted sample size

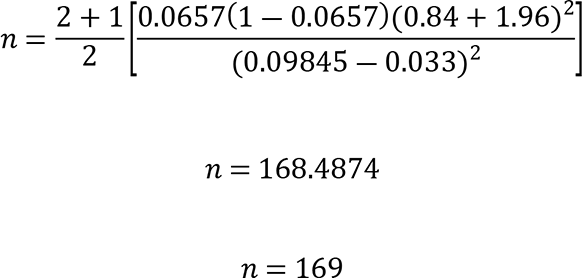

Therefore, the calculated sample size was 169 cases and 338 using the 1:2 ratio, which gave 507. After factoring in 10% additional in case of participant fall out, the total sample size calculated was 558.

### Sampling technique

Sample size per facility based on probability proportional to size sampling after reviewing the previous year records from January to December 2023. The total number of participants enrolled was 635 from all study sites. However, for analysis only 630 underwent analysis while the remainder did not meet the inclusion criteria. Upon enrolment of a case, two controls randomly selected from within the 24 hours shift that a case occurred. The disparity in the number since it was a 1:2 selection is because of exclusion during data cleaning and some entry errors.

### Study variables

The outcome variables include macerated, fresh stillbirths, and early neonatal death. Distal level variables included age, marital status, parity, gravida, weight, BMI, employment status, educational level, distance of place of residence to health facility, and mode of transport. Intermediate level variables were antenatal booking data, number of antenatal contacts, medical history, history of pregnancy loss beyond viability, history of abortion, malaria during pregnancy, Human Immunodeficiency Virus (HIV) status, Tuberculosis during pregnancy, syphilis, alcohol intake during pregnancy, illicit drugs, smoking, folic and ferrous use during pregnancy, malaria treatment during pregnancy, and sickle cell anaemia. Rhesus factor, antepartum haemorrhage, preeclampsia, and eclampsia, preterm delivery, history of low birth weight, and inter-pregnancy interval were the rest. While proximal level variables included gestational age at the time of birth, outcome, apgar score, time lag to Neonatal Intensive Care Unit (NICU), age of early neonatal at the time of demise, sex, gross congenital anomalies, congenital louis, prematurity, low birthweight, and cause of death.

### Data collection tool

A standard interviewer administered questionnaire was set up using the google sheet platform with protected access to the selected data collectors only. Control was exclusive to the PI for daily monitoring and management in real time. After completion of data collection the data set was downloaded to an excel format for screening and cleaning and then coded to construct a do file for execution in Stata version 15.

### Data analysis

Descriptive statistical analyses summarized of proportions and frequency of cases and controls. Univariable analysis determined the crude association between perinatal mortality and independent variables. To demonstrate association with a p-value of <0.05, multivariable logistic regression analysis to show the association. Sequential elimination of predictors checked for strength and significance of the variables of the perinatal deaths followed by discrimination and calibration of the model. Some continuous variables such as age, weight, parity, and gravida introduction into the model as categorical variable was on intuitive from a clinical standpoint. Assessment of the ability to allocate appropriate risk occurred using Hosmer and Lemeshow goodness of fit test. The interpretation of the Hosmer and Lemeshow goodness of fit test was accurate when it yielded a non-significant *p-*value (*p*>0.05). Model stability measurement was by discrimination and differentiation of participants with perinatal death from those without by using area under the receiving characteristics curve. The acceptable area under the curve was 0.7 (20). Strengthening the Reporting of Observational Studies in Epidemiology (STROBE) guideline were followed to report this study (21).

### Ethical considerations

Ethics approval for this study was through the University of Zambia Biomedical Research Ethics Committee (Unzabrec) (referee 3718-2023). This followed clearance by the National Health Research Authority (NHRA) reference number NHRA 000012/16/03/2023. There was Confidentiality and privacy for study participants. The participants’ anonymized credentials and data collected remained on kept on password-controlled gadgets only accessible by the PI.

## Results

### Description of study participants

The total perinatal deaths 210 (cases) and 420 with live births (controls) made the ratio of cases to controls 1:2. The proportion of the perinatal outcomes were early neonatal death 118 (56%), Macerate Stillbirth (MSB) 56 (27 %), and 36 (17%) Fresh Stillbirths (FSB) respectively.

### Participants’ socio-demographic characteristics

From the social-demographic attributes (distal level variables) of the respondents sampled majority of the respondents were between the age of 20-34 years 401 (68.2%). 19 years old and under were 90 (15.3%) and 97 (16.5%) were 35 years of age and above. Mean age for the study population was 27. Married respondents were 478 (77%) and 143 (23%) not married. 487 (85.6%) had parity between 1 to 4 while the grandmultipara were 98 (17.3%). 445 (72.4%) represented the unemployed and 170 (27.6%) unemployed. In terms of religious background, respondents included 182 (31.4%) 120 (20.7%), 207 (35.8%), and 70 (12.1%) catholics, protestants, pentecostal, and other unspecified religious groupings. For education, the bulk of the respondents had only attained secondary level of education 331 (54%), primary 187 (30.5%), and 95 (15.5%) for tertiary. Most of the participants used public transport 415 (67%) while only 98 (15.8%) had personal transport and the others 106 (17.1%) on foot.

### Predictors of perinatal mortality from bivariate analysis

Bivariate analysis exposed several variable associated with perinatal mortality. The outcomes outlined in the table below revealed that the level of education of the mother had a significant association with perinatal mortality (p<0.017). The form of transport used when going to the hospital also had association (on foot) with perinatal mortality (p<0.001). Antenatal Care (ANC) booking (first contact with healthcare provider for ANC) beyond 12 weeks of gestation was statistically significant with p<0.001. Women who had prior history of perinatal death also had a statistically significant p-value (p<0.001). Abortion and Anaemia during pregnancy were found with p <0.018 and p<0.003 respectively. The rest of the independent variables had no statistical significance. The following table 1.0 summarised maternal and socio-demographic characteristics and frequency.

### Unadjusted logistic regression of variables associated with perinatal mortality

According to the univariate and multivariable logistic regression for factors associated with perinatal death in table 5. The odds of experiencing perinatal mortality were almost 2 times more likely to primary education level respondents compared to the tertiary level group (OR=1.998, 95% CI: 1.12-3.56). The findings also indicate that the odds of perinatal mortality to occur were slightly over 3 and almost twice times (OR=3.482, 95% CI: 1.87-6.49) and (OR=1.964, 95% CI: 1.15-3.34) more likely to occur in woman whose mode of transport was on foot and public transport respectively, comparison to those with personal transport to access healthcare services. Previous loss of a baby before birth was almost 5 times more (OR=5.273, 95 CI: 3.31-8.402) likely than those without prior loss a baby before birth. This study also revealed anaemia during pregnancy odds of experiencing perinatal mortality was more than two and half times likely (OR=2.513, 95 CI: 1.34-4.69) in comparison with those without anaemia during pregnancy. History of abortion had three time likely (OR=3.102, 95% CI: 1.16-8.28) to experience perinatal mortality compared to those who had no history of abortion before.

In order to understand the combined impact of factors associated with perinatal mortality and to control for confounding, it required multivariable logistic regression analysis. The above table is the illustration of the final multivariable model that best describes the factors associated with perinatal mortality. Following a thorough analysis and correction for the impact of extraneous variables, three variables stood out as being associated with perinatal mortality. The odds of perinatal mortality occurrence due to first antenatal care (booking) visit beyond 12 weeks of gestation was almost three times greater than that of mothers with early booking (less than 12 weeks) (AOR 2.909, 95% CI: 1.97-4.296), p <0.001. Anaemia during pregnancy was three and half times more associated with perinatal mortality than those without anaemia did in pregnancy (AOR 3.581, 95% CI: 1.72-7.44). Lastly, history of loss of baby before birth before was five times more likely to experience perinatal mortality compared to those who had not (AOR 5.047, 95% CI: 2.99-8.51). Table 2.0 hereafter is a summarized comparison of univariable and multivariable predictors after univariate and then multivariate logistic regression analysis.

**Table 2.0.** Multivariable logistic regression analysis for factors associated with perinatal mortality in the selected study sites.

| Univariate logistic regression |  |  |  | Multivariate logistic regression analysis |  |  |
| --- | --- | --- | --- | --- | --- | --- |
| Predictor variable | Odds Ratio (OR) | 95%CI | p-value | Adjusted Odds Ratio (AOR) | 95% CI | P-value |
| **Educational level** |  |  |  |  |  |  |
| Tertiary | Ref |  |  |  |  |  |
| Primary | 1.998 | (1.12-3.56) | 0.019 | 1.658 | (0.83-3.31) | 0.152 |
| Secondary | 2.161 | (1.26-3.71) | 0.005 | 1.869 | (0.98-3.57) | 0.058 |
| **Mode of Transport** |  |  |  |  |  |  |
| Personal | Ref |  |  |  |  |  |
| Foot | 3.482 | (1.87-6.49) | <0.001 | 2.75 | (1.23-5.39) | **0.012\*** |
| Public | 1.964 | (1.15-3.34) | 0.013 | 1.70 | (0.92-3.16) | 0.091 |
| **ANC Booking** |  |  |  |  |  |  |
| < 12 weeks | Ref |  |  | Ref |  |  |
| > 12 weeks | 3.682 | (2.56-5.30) | <0.001* | 2.909 | (1.97-4.3) | **<0.001\*** |
| **History of Stillbirth** |  |  |  |  |  |  |
| No | Ref |  |  | Ref |  |  |
| Yes | 5.273 | (3.31-8.40) | <0.001* | 5.047 | (2.99-8.51) | **<0.001\*** |
| **History of abortion** |  |  |  |  |  |  |
| No | Ref |  |  |  |  |  |
| Yes | 3.102 | (1.16-8.28) | 0.024 | 0.858 | (0.24-3.06) | 0.814 |
| **Anaemia during pregnancy** |  |  |  |  |  |  |
| No | Ref |  |  | Ref |  |  |
| Yes | 2.513 | (1.34-4.69) | 0.004 | 3.581 | (1.72-7.44) | **0.001\*** |

## Discussion

The aim of this study was to assess the predictors of perinatal mortality in the selected study sites of Lusaka, the capital city of Zambia. Mode of transport that the women utilized when accessing healthcare during pregnancy, timing of antenatal care booking, history of abortion, stillbirth, and experiencing anaemia during pregnancy were strong predictors of PMR.

We found that the proportions of fresh and macerated stillbirths (17% and 27% respectively) as well as neonatal deaths (56%) remained high. This is in line with some previous studies that reported results of a similar nature (22). Shelke, Shivkumar and Chaurasia (23) also reported ratios of more macerated to fresh stillbirths. The authors attributed this to be a reflection of the quality of prenatal and obstetric care, with higher fresh to macerated ratios implying poorer care. For macerated stillbirths utero-foetal demise occurs without onset of labour within 8 hours from the time of demise (24). Maceration onset can range from 6 – 12 hours. Factors that contribute to macerated stillbirths include intrauterine growth restriction, infections, and placental lesions, while a large proportion of these deaths remains unexplained (25). Early neonatal deaths also tend to be more because their attributed risk to mortality are a mix of low birthweight, complications that need neonatal intensive care and maternal complications (26, 27). Birth asphyxia, sepsis, prematurity, respiratory illnesses, and other unspecified illnesses have also been reported as the major risks of early neonatal death according to a study done in Ethiopia (28).

Mode of transport was a significant predictor of PMR. Women who walked to the facility to seek medical care during pregnancy has three times more likely to suffer perinatal demise than those who have personal transport. We also found that among those who used public transport, the risk was almost two times higher. This finding is in tandem with a previous study done in Zambia and Uganda that reported motorized vs non-motorized means of transport to have significant difference statistically (29). This is because women in the poorest quintile in Zambia compared to those in the wealthiest quintile took an hour more to seek obstetric services. The study demonstrated that women who used motorized means of transport had less risk of perinatal mortality compared to those who did not. One of the possible reasons may be due to lack of women empowerment, including economical, for early decision-making autonomy, poor physical accessibility to healthcare facilities and lack of access to health education regarding preparedness when labour commences (30). Another plausible reason is according the study in Uganda and Zambia by Sacks (29) that found that women who travelled by foot were more likely to only access basic emergency obstetric and neonatal care (BEmONC) facilities. Their counterpart, who used motorized transport were more likely to deliver at comprehensive emergency obstetrics and neonatal care facilities (CEmONC). CEmONC facility services allow women with complications during pregnancy to have access to better quality obstetrics care than what is obtainable at BEmONC facilities (31)..

Early antenatal care booking plays a key role in detecting and treating some complications of pregnancy and creates a good basis for appropriate management during delivery and after (32). The WHO ANC model recommends the first ANC contact scheduled to take place in the first trimester, up to 12 weeks of gestation (33). In our study, more than 70% of the respondents booked late (>12 weeks gestation) for ANC. Moreover, our study demonstrated that late ANC initiation had three times the risk of perinatal mortality compared to those who started early (<12 weeks gestation) as prescribed by the WHO ANC guidelines for positive pregnancy experience (33). This experience is supported by other studies too that reported on early antenatal care failure resulting in potential complications during pregnancy, delivery, and puerperium that inadvertently increase risk of perinatal mortality (34, 35).

Women who had had a history abortion before also had three time more at risk of having a perinatal death compare to the women who had not. A recent study on stillbirths determinants reported similar findings of higher risk of stillbirth among those with prior experience of stillbirth (36). This is also in line with another similar study by Dube and Lavender (37) in Zimbabwe that reported similar findings. History of abortion in this study demonstrated statistical significance as predictor of perinatal mortality. The finding is consistent with a study done in Ethiopia on the effects of previous stillbirth or abortion on subsequent pregnancies and infants increased the risk of infections (38), which increase the risk of perinatal mortality (39). In furtherance to this, we also found that having a history of stillbirth had five times higher risk of experiencing perinatal mortality compared to those who had never. Most stillbirths lack of explanation on causality especially due to the low rates of post-mortem investigations availability (40). This makes experiencing stillbirth is a difficult conversation not only to the parents but to professionals too as they endeavour to obtain consent for investigations to try ascertain the cause of death (41) The risk of having a perinatal mortality was two and half times higher among women with anaemia. Anaemia in pregnancy has been associated with higher rates of small for gestational age and preterm birth, and these pose a high risk factor for perinatal mortality (42). Small for gestational age, preterm birth and low birth weight demonstrated high risk for perinatal mortality in previous studies done in Zambia (17, 43). However, in this study, we failed to demonstrate that small for gestational age and preterm birth were risk factors for perinatal mortality.

## Strengths and Limitations

The study was able to investigate multiple predictors simultaneously. In this investigation’s setting, the sample collected from the population is representative of Lusaka as all the major hospitals contributed to the total sample. This makes the findings generalizable. The identification of significant predictors provides evidence to not only public health and policy makers but to clinical practice too. Some limitations could have affected the findings for this case-control study. The first notable is lack of completeness of some information during data collection due to inconsistencies with medical record keeping. Stating or recording cause of death is difficult in stillbirths because investigations are not routine in an endeavour to assign cause of death at least.

## Data Availability

We will make full availability and without restriction all data underlying the findings if requested

## Acknowledgements

I would like to thank all my supervisors for the guidance provided to complete this study. I also would like to acknowledge the immense contribution the study participants for accepting to be part of the project.

## Authors’ contributions

MM: conceptualization of the study, study design, data collection, data cleaning, data processing and analysis, and manuscript writing. PK: supervision and methodology. CJ: supervision and contextualization of the project. MM: conceptualization and contextualization of the study. BV: overall content supervision, contextualization of the study, and methodology. All authors have read and approved the final version of the manuscript.

